# The Impact of Sustained LDL-C Elevation on Plaque Changes: Primary Coronary plaque progression results from the Keto CTA Study

**DOI:** 10.64898/2026.01.15.26343955

**Authors:** Matthew J. Budoff, April Kinninger, Venkat S. Manubolu, Nicholas G. Norwitz, David Feldman A.A., Adrian Soto-Mota

## Abstract

**Background:** Carbohydrate-restricted diets are gaining popularity, including among lean individuals. In these populations, a lipid phenotype often emerges comprising elevated LDL cholesterol (LDL-C), alongside elevated HDL-C and low triglycerides, termed the *lean mass hyper-responder* (LMHR).

**Objective:** To evaluate one-year coronary plaque progression in LMHRs and near-LMHRs.

**Methods:** This prospective study followed 100 participants who developed the triad of high LDL-C, high HDL-C, and low triglycerides after adopting a ketogenic diet over one year. Coronary plaque changes were assessed using coronary CT angiography and analyzed using the prespecified QAngio^®^ methodology (Leiden, the Netherlands), with AI-enabled coronary plaque analysis (AI-CPA; HeartFlow^®^ Inc., Mountain View, CA) used as an independent, blinded confirmatory analysis. Plaque burden and plaque progression predictors were examined using linear regression.

**Results:** All 100 participants with elevated LDL-C and a mean BMI of 22.5 ± 2.7 kg/m2 completed the study. At baseline, 57 (57%) had zero CAC. After follow-up, most participants remained with low-risk plaque burden markers: 81% of participants had a CAC score <100, and 54% had a CAC of 0. The median increase in non-calcified plaque volume was 5.6 mm³ (37% relative increase). Notably, 15% of participants exhibited plaque regression despite sustaining elevated LDL-C (mean 242 mg/dL) and ApoB (mean 180 mg/dL). Additionally, 78% had percent atheroma volume (PAV) below the high-risk threshold of 2.6%, and 93% had total plaque volume (TPV) below the high-risk threshold of 254 mm³. Baseline plaque metrics were consistently predictive of plaque progression. By contrast, neither ApoB levels nor cumulative LDL-C exposure predicted plaque progression in this population of LMHR and near-LMHR individuals.

**Conclusion:** These findings suggest that over one year, progression was modest and heterogeneous in this population, with baseline coronary plaque emerging as the strongest predictor of subsequent plaque progression in LMHRs, whereas traditional lipid markers such as ApoB and LDL are not.

## INTRODUCTION

Carbohydrate-restricted diets (CRD), including very low-carbohydrate ketogenic diets (KD), are becoming increasingly popular for a wide variety of medical reasons^1–3^ or personal preference. While this diet is typically higher in saturated fat, most individuals on CRD do not experience sustained increases in LDL-C^4^.

However, leaner metabolically healthy individuals commonly exhibit increases in LDL-C and ApoB, in conjunction with increases in HDL-C and low triglycerides^5^. This phenotype—termed Lean Mass Hyper-Responder (LMHR)—is defined by the combination of LDL-C ≥ 200 mg/dL, HDL-C ≥ 80 mg/dL, and triglycerides ≤ 70 mg/dL. While the precise mechanisms driving this phenotype are still a matter of debate and active investigation^6, 7^., it has been established that the LMHR phenotype is more strongly associated with lower body mass index (BMI) and adiposity^8^ as well as higher energy expenditure^9^ than saturated fat intake attributed to CRD and KD.

Given the sometimes extreme increases in LDL-C and ApoB in these LMHR and near-LMHR subjects, and considering the peculiarities of their specific phenotype–a lifestyle-induced and readily reversible increase in LDL-C and ApoB that occurs particularly in lean people on CRD or KD – an obvious question arises: what is the cardiovascular risk associated with this phenotype?

It’s widely accepted that ApoB-containing lipoprotein particles are part of the causal cascade of atherosclerosis^10^, and most (if not all) available Cardiovascular Disease Risk (CVDr) methods consider LDL-C (or other ApoB surrogate markers such as non-HDL) as one of their predictors^11^. However, the fact that the LMHR profile is distinct from other populations with elevated LDL-C and ApoB creates an interpretive challenge: it is methodologically inappropriate to assume that CVDr assessments typically found in the general population necessarily generalize to the LMHR phenotype.

For example, it is well-documented that persons with high HDL-C and low triglycerides have low cardiovascular risk, even if they have above “optimal” LDL-C^12^. And even studies in symptomatic individuals with very high LDL-C (190 mg/dL or higher) have shown that cardiovascular risk may be low when other low-risk predictors are present (such as a Coronary Artery Calcium [CAC] score = 0)^13^. In that large observational study, absence of CAC was a prevalent finding (438 of 948 [46.2%] in patients with LDL-C levels of at least 190 mg/dL) and associated with no detectable plaque on CT angiography in 338 of 438 (77.2%).^13^

As CRD adoption increases among lean and insulin-sensitive individuals, prospective evidence on the cardiovascular risk implications of high LDL-C and ApoB in the absence of other risk factors is largely absent from the literature, and only a handful of studies have made use of modern angiotomographic techniques.^14,15^ Therefore, we aimed to measure longitudinally plaque progression and its predictors in people with an LMHR phenotype using CCTA.

## METHODS

### Design and Study Population

This study (NCT05733325) was conducted in compliance with the principles of the Declaration of Helsinki and relevant local laws and regulations. Written informed consent was obtained in all cases. Potential participants were pre-screened based on the full eligibility criteria and the availability of supporting medical documentation to confirm serum markers. The inclusion and exclusion criteria, as well as the participants’ workup during the study, have been summarised in previous publications^16^.

### Dietary and Metabolic Measurements

All participants were asked to participate for one year, and their adherence was assessed through three dietary recalls. Dietary intake was assessed using the Automated Self-Administered 24-Hour (ASA24) Dietary Assessment Tool. Blood β-hydroxybutyrate (βHB) was measured using a handheld monitor (KetoMojo, Napa, CA)^16^. Ten-year Cardiovascular risk was calculated with *CVrisk::chd_10y_mesa*, which incorporates CAC values.

All CCTA scans were performed at baseline and one year after at The Lundquist Institute for Biomedical Innovation at Harbour-UCLA Medical Centre using the 256-slice multidetector computed tomography (GE Revolution; General Electric) and were blindly read by Level 3 cardiac CT readers. A nonenhanced ECG-gated CAC scan was also performed before each CCTA.

The prespecified primary method for plaque volume quantification was QAngio^®^ (Medis, Leiden, the Netherlands)^16^, and all scans were evaluated by experienced readers unaware of the identity and sequence of scans. This software automatically detects lumen and vessel border contours, with manual correction by expert readers in areas of misregistration. Second, AI-CPA (Heartflow^®^ Inc., Mountain View, CA) was used as a sensitivity analysis.

The primary outcome was the observed per-subject rate of change in the coronary atherosclerotic plaque burden during the observation period^16^. Noncalcified plaque volume change was the primary outcome, with changes in other plaque metrics, including total plaque, low attenuation, fibro-fatty, fibrous, calcified plaque volumes, percent atheroma volume (PAV),^17^ and CAC measures serving as secondary outcomes. Additionally, the presence, incidence, and progression of other CCTA metrics and CAC scores, as both dichotomous and continuous outcomes, were pre-registered, as well as exploratory analyses and comparisons with different subjects and studies^18^.

Finally, for the dichotomous outcome comparisons on high-risk plaque burden markers, we used the cut-off points established in other studies using CCTA.^19^ High-risk plaque burden markers were defined as Low Attenuation Plaque (LAP) of 4% or higher^20^, PAV of 2.6% or higher, and Total Plaque Volume of 254 mm^3^ or higher^21^. Rapid plaque progression was defined as an annual change of more than 1% in PAV^19,20^.

### Statistical Analyses

Data are presented using the median with 25^th^-75^th^ percentiles and the mean with standard deviation (SD) for continuous variables, and counts (percentages) for categorical variables. Data cleaning and statistical analyses were done using R version 4.4 (R Core Team [2024]. R Foundation for Statistical Computing, Vienna, Austria). Unless specified otherwise, all functions use default settings. Linear models were univariable and analyzed using *the stats::lm function, and all linear model assumptions were corroborated using* the R function *performance::check_model.* LDL-C exposure was defined as baseline LDL-C plus the measured change during follow-up.

Since there are no comparable CCTA-based quantitative plaque progression studies with similar subjects, our sample size calculation was necessarily based on limited available data. We examined plaque changes in placebo groups of clinical trials using cardiac CT angiography, which showed substantial variability (20 to 30 ± 40 to 80 mm³). Without a priori knowledge of the baseline plaque burden or progression patterns in the LMHR phenotype, we designed the study with 100 subjects (assuming 15% dropout) to provide adequate power to detect meaningful changes and to characterize the distribution of plaque progression in this population. Although it was not part of the original sample calculation, we estimated the available statistical power to observe an association between baseline LDL-C and plaque changes. According to the available literature, this correlation between LDL-C and plaque changes has ranged between 0.4 and 0.6^22,23^, with an explained variability of between 0.1 and 0.4. Using GPower version 3.1, we estimated that ∼80 participants would be needed to observe an association as low as 0.3 and an R^2^ as low as 0.09 (**Supplemental Material**).

Bayes factors were calculated using BayesFactor::regressionBF with default settings. To allow for potentially large effect sizes—consistent with prior literature on LDL-C and plaque progression—an rscale value of ∼0.8 was used. For their reporting, Jeffrey’s scale was used to establish the magnitude that qualified as weak, moderate, or strong evidence in favor of or against the null hypothesis.^24^

To further support that the association between LDL-C exposure and plaque change was negligible, we conducted a two one-sided test (TOST) procedure for hypothesis equivalence on the regression slope with the function parameters::equivalence_test using default settings (rule = “classic”, and CI = 0.9). Equivalence bounds were set a priori at ±0.05 for detecting even a clinically negligible regression coefficient. Finally, sensitivity analyses included robust linear regression (using MASS::rlm and sfsmisc::f.robftest).

## RESULTS

At the start of the study, most participants had been on a KD for more than 4 years (mean KD duration 4.5 years), and, except for high ApoB, most participants presented markers of low cardiovascular risk, with mean BMI (22.5 ± 2.7 kg/m^2^), mean systolic blood pressure (121.8 ± 21.1 mmHg), and low serum triglycerides (66.7 ± 30.4 mg/dL). Additionally, 83% exhibited low-risk CAC <100 (**Table 1**). Using QAngio, 85% had plaque at baseline, median 47.5 (IQR 15.5:137.8).

After a one-year follow-up and more than five years of sustained high LDL-C and ApoB, most participants exhibited low absolute plaque volumes, as indicated by NCPV and TPV. Regarding PAV and LAP, individual change also remained low (**Table 2 and Figure 1**). The median plaque progression rate of change was 37% in relative terms (calculated as absolute change divided by baseline value); however, this needs to be interpreted in the context of low mean and median baseline plaque burden, which can inflate relative progression percentages due to a low denominator **(Table 1)**. In absolute terms, the median change in NCPV was 5.6 mm³ (**Figure 1A**).

**Figure 1.**
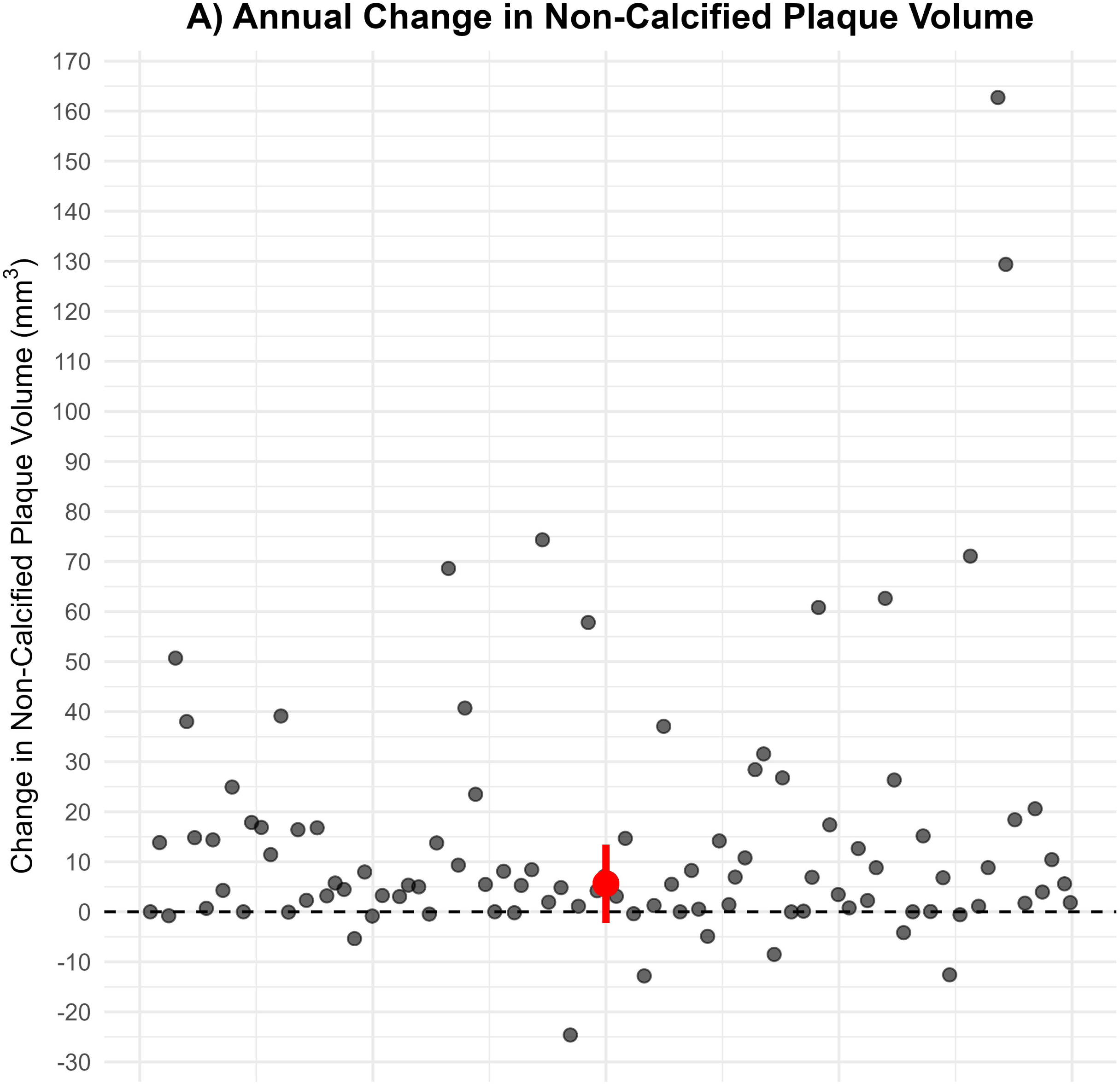
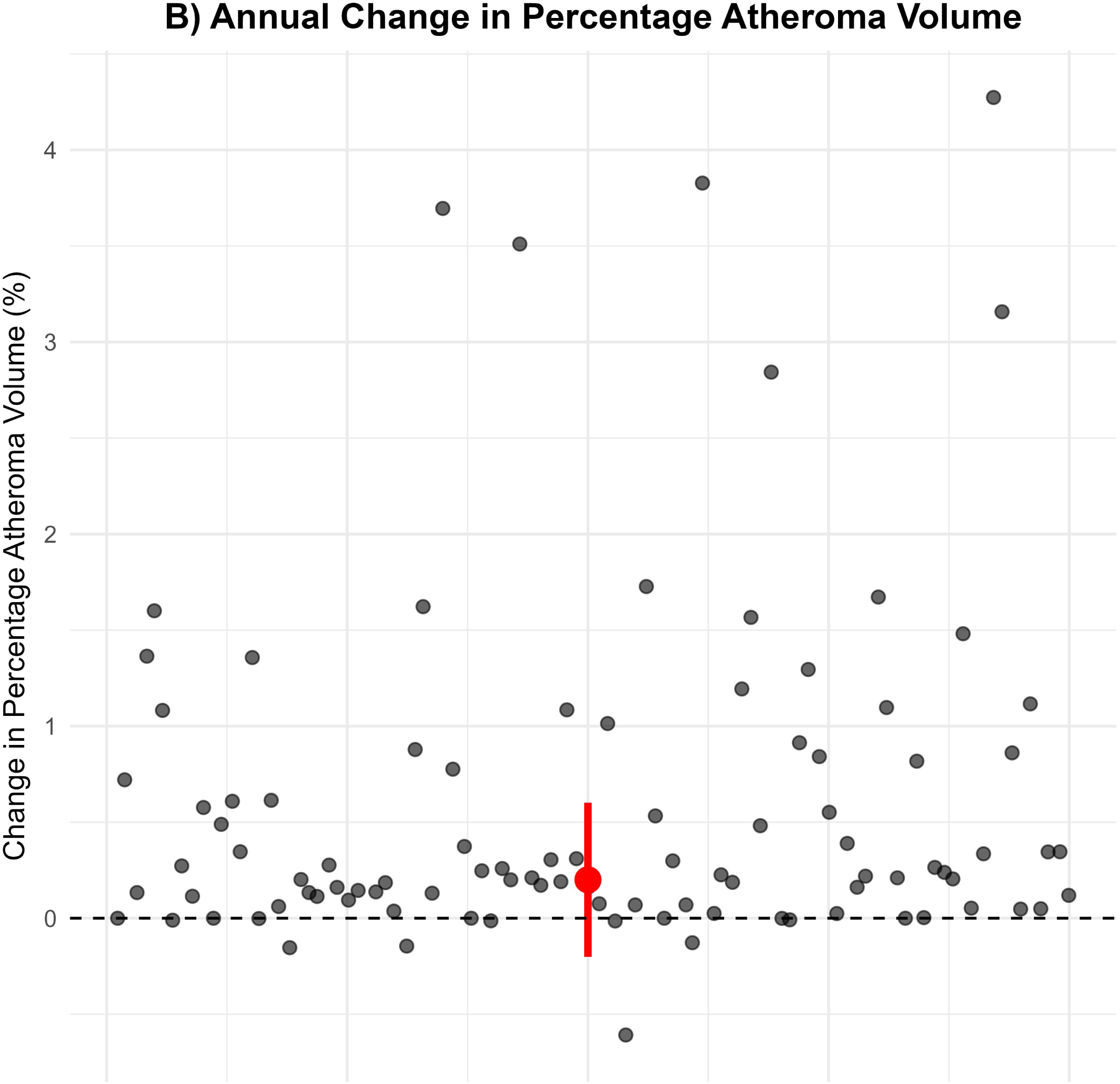
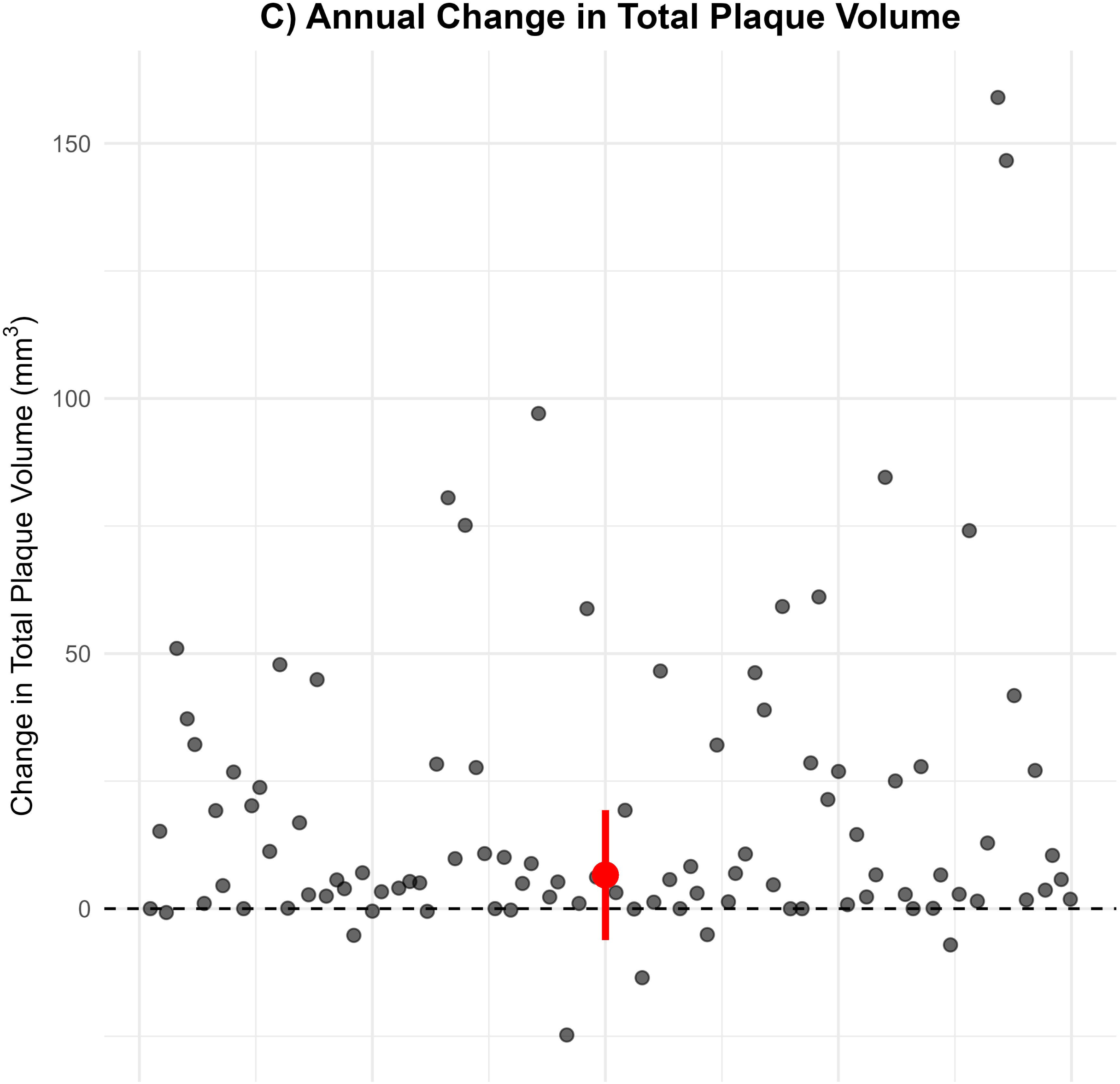
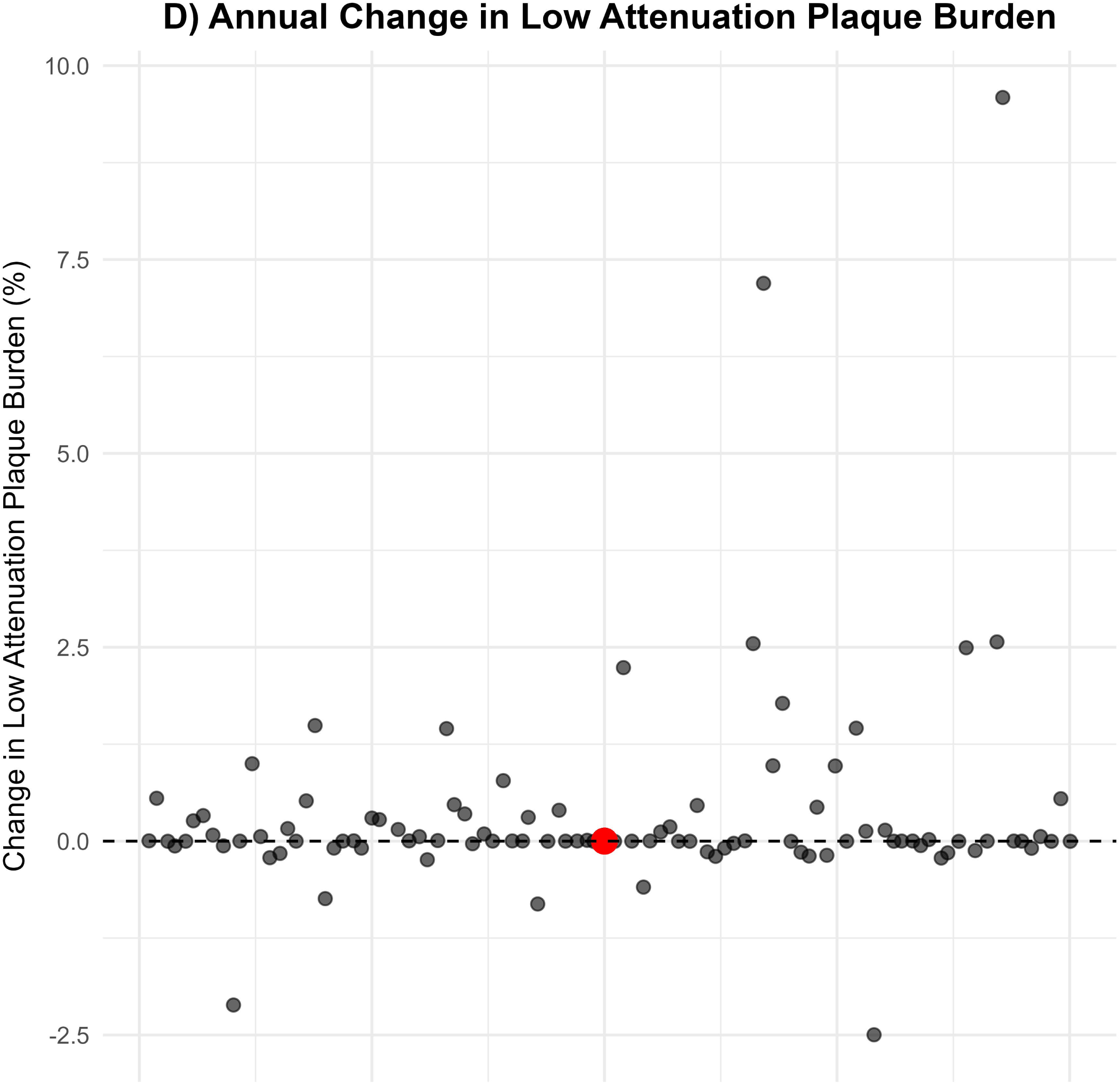
Annual Change in Primary and Secondary Outcomes with QAngio^®^ (pre-specified CCTA method). The red dot illustrates the respective median annual change and its whiskers, its interquartile range.

Fifteen of 99 participants (15%) exhibited plaque regression in the pre-registered QAngio^®^ analysis, as measured by a decrease in NCPV from baseline to follow-up (mean change: -5.1 mm³, range: -24.6 to -0.02 mm³). Additionally, 33 of 95 participants (35%) exhibited regression in the blinded HeartFlow^®^ analysis (mean change: -24.4 mm³, range: -236.0 to -2.0 mm³). Seven participants demonstrated regression with both methods and notably stronger regression magnitudes, suggesting these changes represent robust, detectable plaque reduction.

Among the broader cohort, after follow-up, most participants exhibited low cardiovascular risk plaque burden predictors, with 80% exhibiting CAC<100 and 54% CAC = 0. Moreover, 78% had a PAV of 2.6% or lower, and 93% had a TPV of 254 mm³ or lower. Conversely, only a minority of participants exhibited high-risk plaque metrics, with 21% having a PAV annual change of 1% or higher and only 4% exhibiting an LAP of 4% or higher (**Table 2**). HeartFlow^®^ analyses yielded comparable results (**Table 3 and Figure 2**). One revascularization event was confirmed in 1 subject between scans.

**Figure 2.**
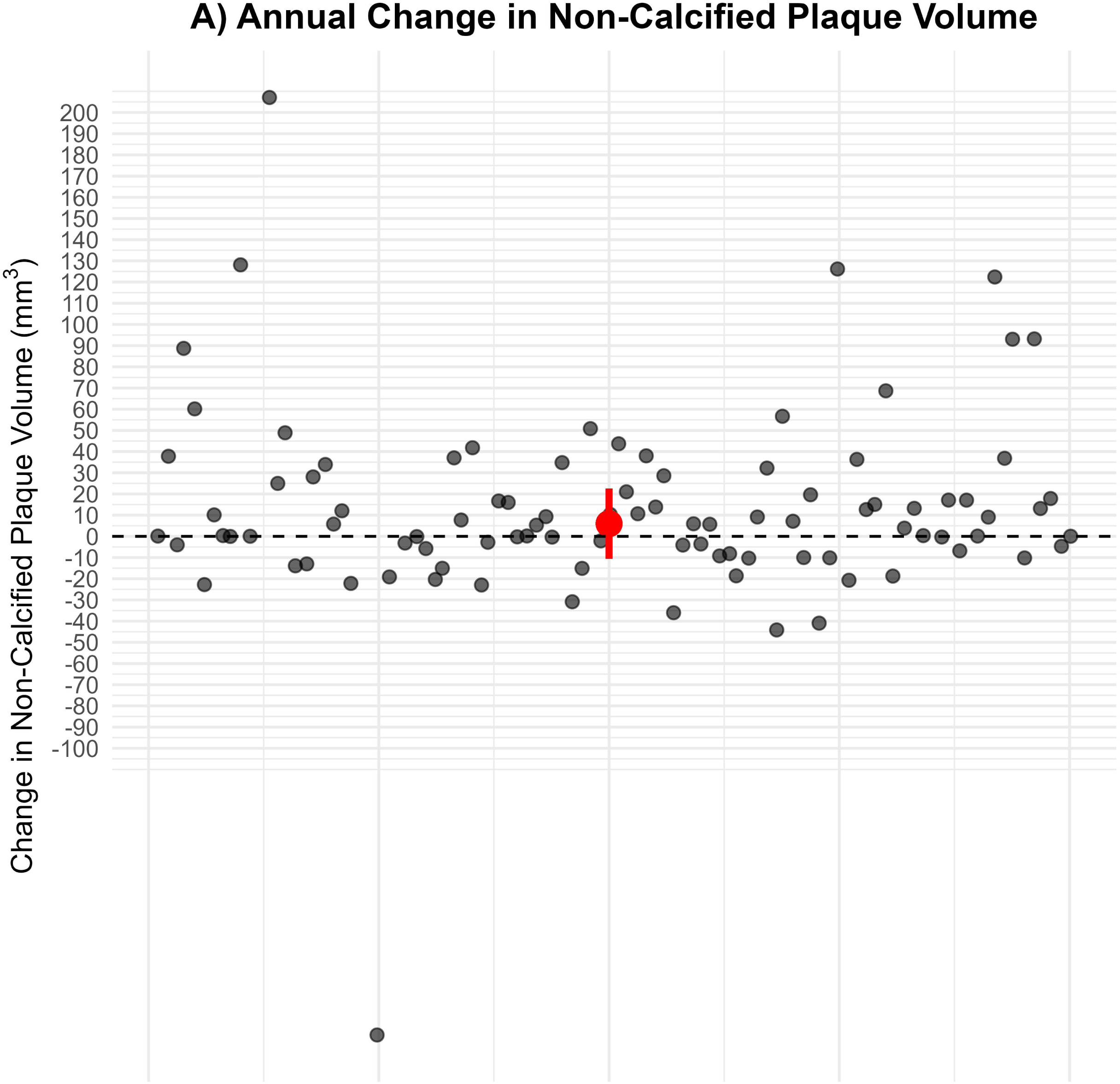
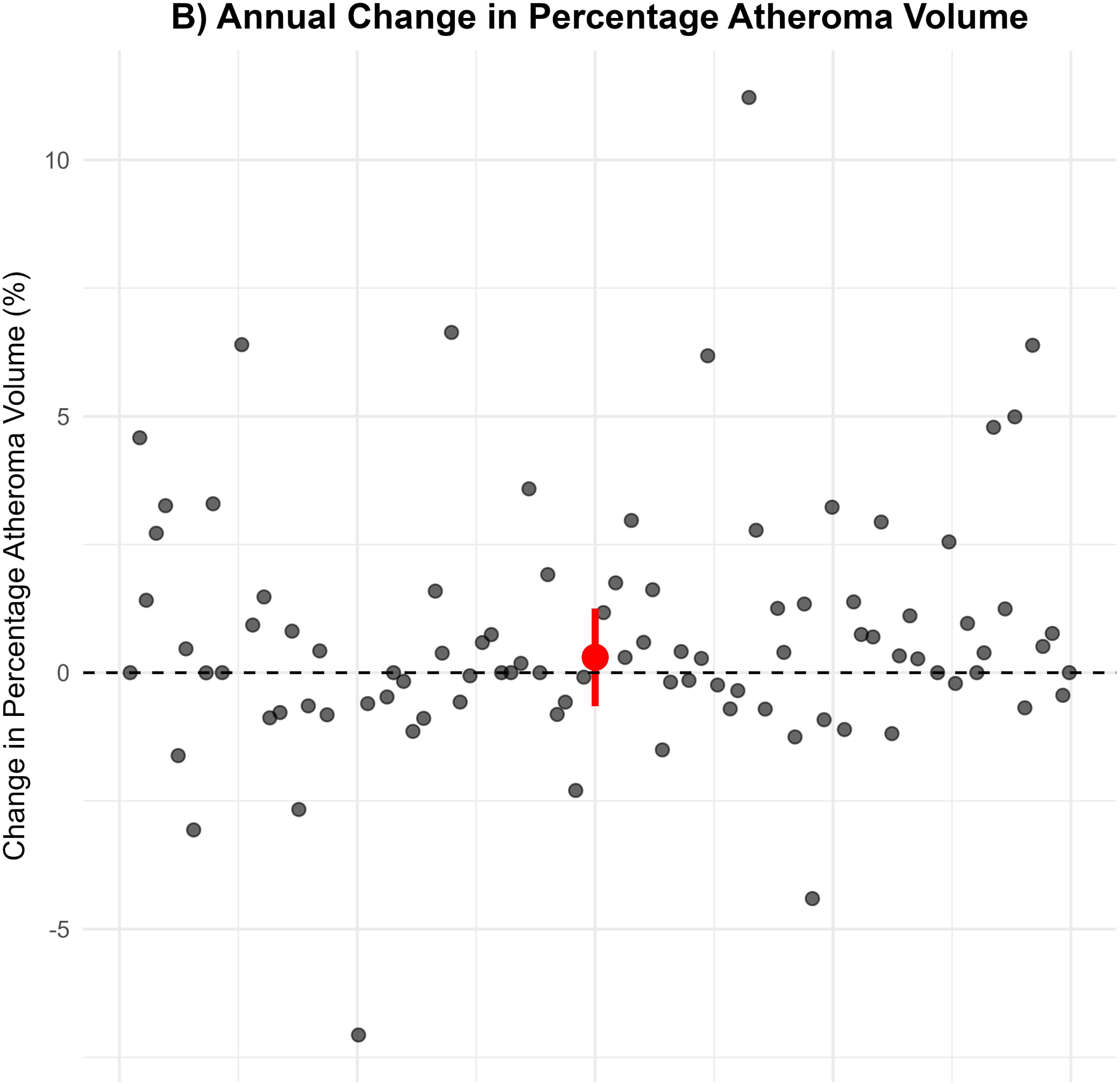
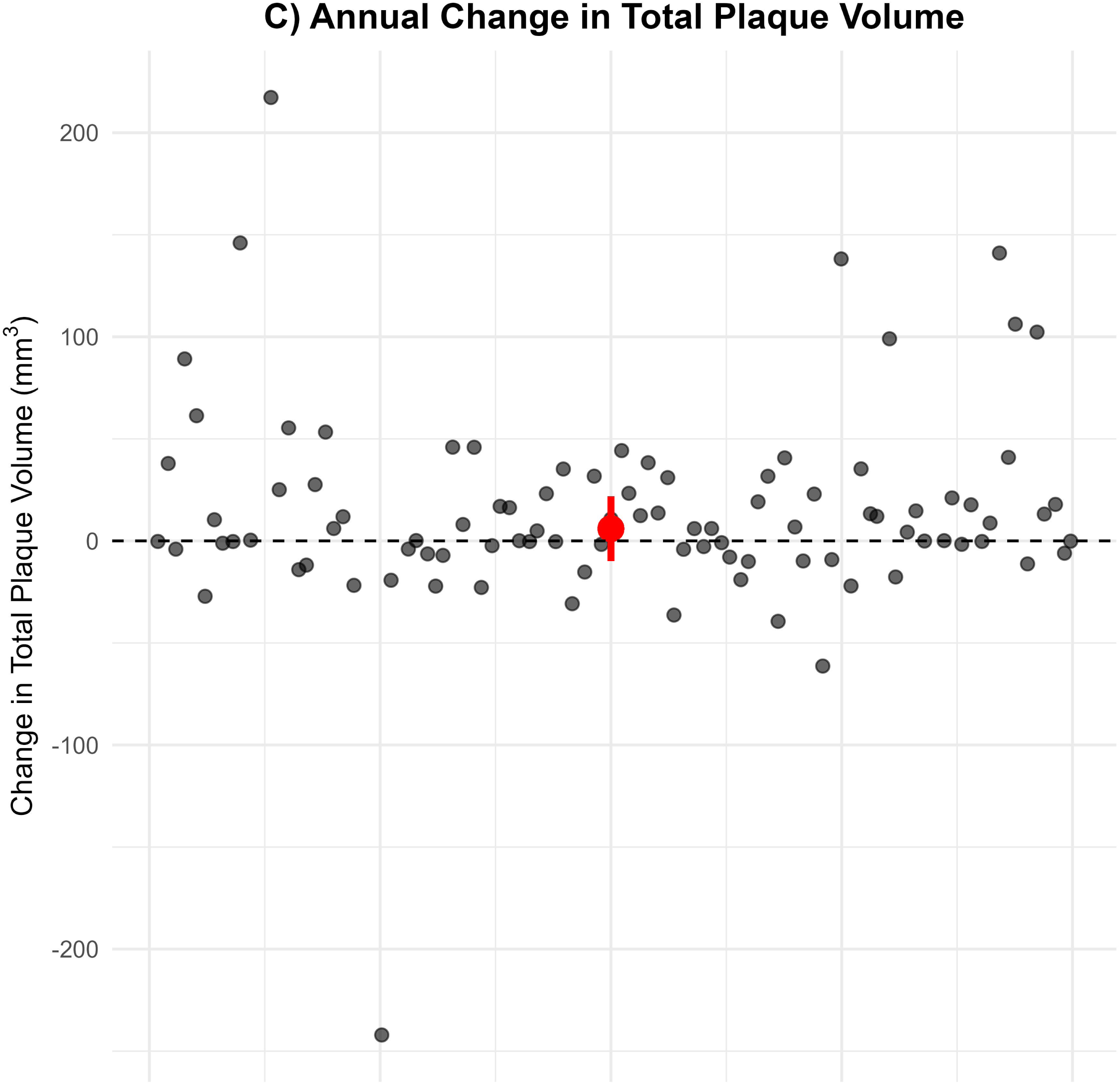
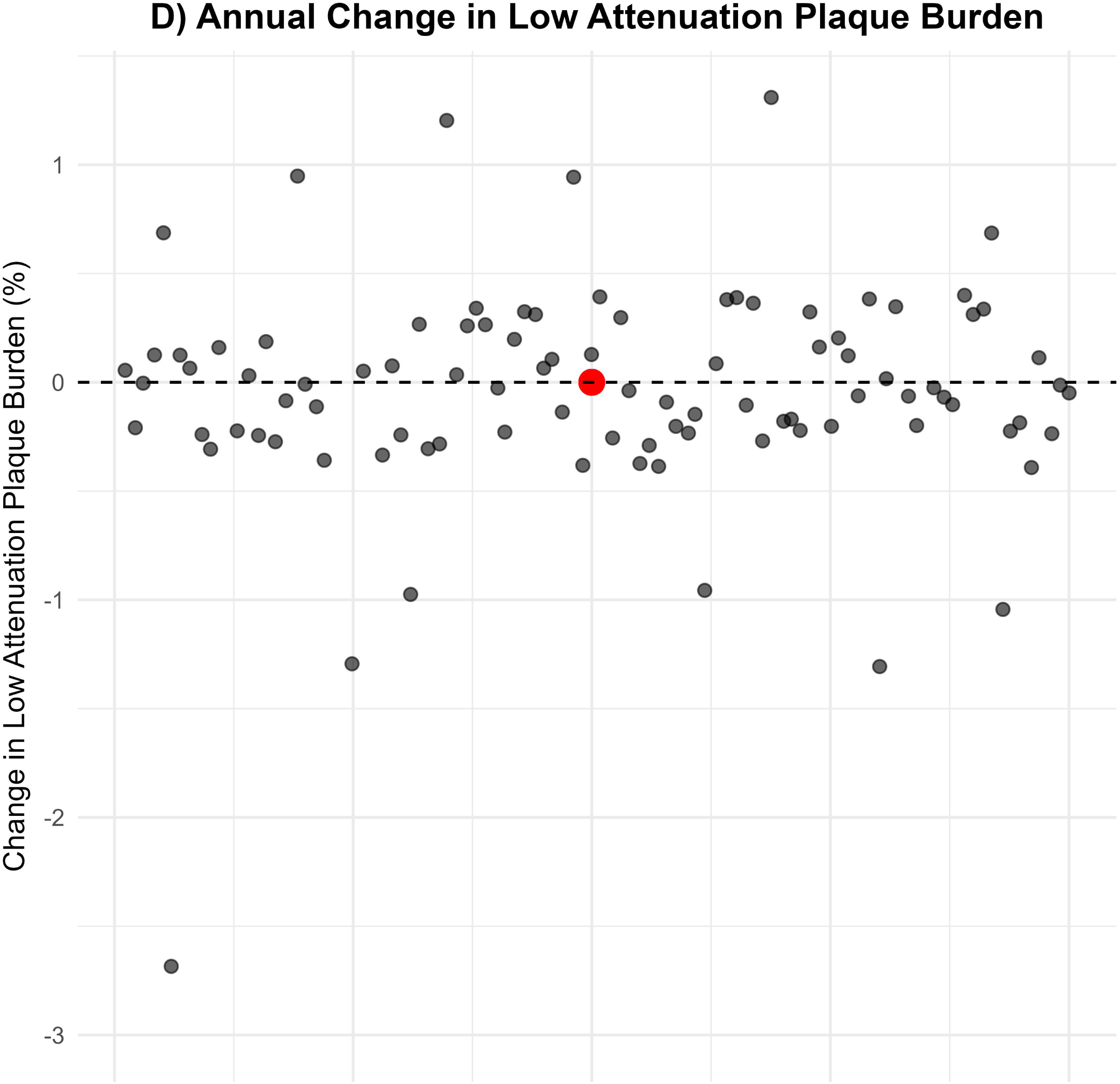
Annual Change in Primary and Secondary Outcomes with HeartFlow^®^ (sensitivity analysis). The red dot illustrates the respective median annual change and its whiskers, its interquartile range.

Importantly, all baseline plaque metrics were strongly predictive of all forms of plaque progression. But neither ApoB exposure during the study nor total LDL-C exposure was predictive of changes in plaque (**Supplemental Material**). For lipid exposure models, the reported values come from unadjusted models. However, baseline plaque-adjusted models were evaluated for all of them and yielded consistent results.

On the other hand, Bayes Factors consistently provided a strong association in favor of the null hypothesis, and the only significant predictor of plaque progression was existing baseline plaque. Further in support of the null hypothesis, all TOST procedures on null association regressions supported that the regression slopes were statistically equivalent to zero with a p < .001, 90% CI [0.00, 0.00]. Even TOST procedures with stricter confidence interval bounds of 98% (equivalent to an alpha = 0.01) yielded similar results. While the linear models met the necessary assumptions for linear regression after visual inspection, robust regression models yielded similar results.

## DISCUSSION

It stands to reason that the relationship between LDL-C and ApoB exposure and cardiovascular plaque progression will vary among individuals. This relationship can be influenced by many factors, including genetics, comorbidities, and the pathophysiology underlying the aetiology of elevated circulating lipoproteins.

Our observations support the notion that the physiological differences between LMHRs on CRD and KD versus other populations exhibiting LDL-C elevations create a distinct cardiovascular risk profile, with 15% of participants showing plaque regression over one year despite elevated LDL-C levels. These findings provide additional context for understanding cardiovascular risk markers in this population and demonstrate the association between CAC and CCTA measures and plaque burden.

We have previously published the baseline plaque variables, demonstrating that despite markedly higher LDL-C than the matched control group, plaque volumes were similar.^18^ It is important to note that these baseline plaque measurements occurred after a mean of 4.5 years on a ketogenic diet. Since baseline existing plaque and baseline cardiovascular risk were associated with plaque changes, these results provide some cardiovascular nuance for patients with low CAC scores who exhibited low final plaque burden and plaque progression metrics, which is consistent with what has been observed in other populations. By QAngio^®^, 21% had significant progression, as defined as a PAV annual change of ≥1%, and 29% by AI-CPA, most notable for those with existing baseline plaque. These findings highlight the relevance of baseline plaque variables (CAC and/or CT angiography) when interpreting plaque progression in individuals following a ketogenic diet and are consistent with the position that cardiac imaging is a superior clinical risk stratified than traditional lipid measurements.

Thus, these results support the clinical utility of CAC (which has extensive evidence supporting its utility for individualized benefit–risk assessment.^25,26^) and CCTA measurements and emphasize the need for developing more accessible and useful CVDr prediction tools for LMHR and near-LMHR individuals.

To our knowledge, this study is among the first to include two distinct quantitative CCTA plaque analyses applied across all scans. Additional strengths of our study include: Dietary adherence corroborated with daily βHB data, all participants completing their follow-up, and strong representation of different CVDr tiers. Although the study was designed to include participants with high LDL-C, the actual LDL-C values observed span a wide range (Table 1), which allows for contrasts when assessing plaque changes. We have published the reproducibility of these QAngio^®^ methods in our laboratory, and for noncalcified plaque volume, the intraclass correlations (ICCs) and coefficients of variation were 7.8%, the interobserver ICC was 0.99, the interobserver CV was 19.9%, and the interobserver ICC was 0.95.^27^

Moreover, the fact that Bayes Factors converge towards 1 when there is low statistical power, along with the power assessments included in the supplemental material, reassures us that the null association between plaque changes and LDL-C exposure is not due to low statistical power. Along the same lines, TOST procedures are also conservative under low statistical power scenarios, ensuring that they will not conclude there is equivalence unless there is sufficient precision to exclude meaningful effects confidently.

Further, the QAngio^®^ analysis aligns with the AI-CPA analysis. This AI-CPA accuracy has been validated with intravascular ultrasound in multiple studies, with a validated staging system based on plaque volumes.^28–30^

Our study does have limitations. There was no control group without the LMHR phenotype, although a previously published matched analysis found comparable plaque burden in people with much lower LDL-C blood levels^18^. Importantly, plaque quantification methods were not trained to look for zero or little plaque; there could be significant noise and variability in those with minimal baseline plaque. Additionally, CVDr is focused on major adverse cardiac events (MACE) and mortality, rather than plaque progression. It is reasonable to assume, however, that MACE is a direct consequence of plaque progression, as has been shown in both intravascular ultrasound and CCTA studies.^26^ More prospective research with sufficiently long follow-up times to allow for an adequate number of events to occur is needed to confirm the high- or low-risk plaque profile of individual participants.

## CONCLUSIONS

Despite markedly elevated LDL-C and ApoB levels and an average KD duration of exceeding 5.5 years, most LMHR participants maintained low cardiovascular risk markers throughout the study. Baseline plaque metrics were strong and consistent predictors of subsequent plaque progression, whereas neither ApoB nor cumulative LDL-C exposure was predictive of plaque progression. These findings underscore the importance of cardiac imaging tools such as CAC, CCTA, and AI-CPA for accurately assessing cardiovascular risk and for guiding individualized risk–benefit assessments in clinical contexts where LDL-C and ApoB are elevated.

## Supporting information

Main Tables

## Data Availability

The values from all 100 participants for both visits are already publicly available. Please address all further data access requests to Dr Matthew Budoff.

https://citizensciencefoundation.org/keto-cta/

## DISCLOSURES

- **Acknowledgements**: We thank the participants who made this research possible.

- **Data availability:** Please address all data access requests to Dr Matthew Budoff.

- **Conflict of Interest Disclosures:** NGN is a coauthor of a Mediterranean low-carbohydrate-diet cookbook; he donates all royalty payments to nutrition research and education. DF receives financial contributions from membership (e.g., through Patreon) for continued research and is a partner in Own Your Labs LLC. All other authors report no conflicts of interest.

- **Funding/Support:** This study was funded by the Citizen Science Foundation, 7320 S Rainbow Blvd, #102-182, Las Vegas, NV, United States.

## Authors’ contributions

- *Concept and design:* MB, DF, AK, NGN, ASM.

- *Acquisition of data:* MB, VSM, AK.

- *Drafting of the manuscript:* ASM, NGN, DF.

- *Critical revision of the manuscript for important intellectual content:* All authors.

- *Statistical analysis:* ASM, AK.

- *Obtained funding:* DF.

- *Administrative, technical, or material support:* MB, AK.

- *Supervision:* MB, DF.

All authors contributed to the data interpretation and have read and approved the manuscript. No AI-assisted technologies were used.

## ABBREVIATIONS

AI-CPA: Artificial Intelligence-enabled quantitative coronary plaque analysis
ApoB: Apolipoprotein B
BMI: Body Mass Index
βHB: β-Hydroxybutyrate
CAC: Coronary Artery Calcium
CCTA: Coronary Computed Tomography Angiography
CRD: Carbohydrate Restricted Diet
CRP: C-Reactive Protein
HDL-C: High-Density Lipoprotein Cholesterol
KD: Ketogenic Diet
LAP: Low Attenuation Plaque
LDL-C: Low-Density Lipoprotein Cholesterol
LEM: Lipid Energy Model
LMHR: Lean Mass Hyper-Responder
MESAc: Multi-Ethnic Study of Atherosclerosis calculator
NCPV: Noncalcified Plaque Volume
PAV: Percentage Atheroma Volume
TPS: Total Plaque Score
TPV: Total Plaque Volume

## REFERENCES

1. Sethi S, Wakeham D, Ketter T, Hooshmand F, Bjornstad J, Richards B, Westman E, Krauss RM, Saslow L. Ketogenic Diet Intervention on Metabolic and Psychiatric Health in Bipolar and Schizophrenia: A Pilot Trial. Psychiatry Res 2024;335:115866.

2. Phillips MCL, Deprez LM, Mortimer GMN, Murtagh DKJ, McCoy S, Mylchreest R, Gilbertson LJ, Clark KM, Simpson P V., McManus EJ, Oh JE, Yadavaraj S, King VM, Pillai A, Romero-Ferrando B, Brinkhuis M, Copeland BM, Samad S, Liao S, Schepel JAC. Randomized crossover trial of a modified ketogenic diet in Alzheimer’s disease. Alzheimers Res Ther 2021;13:1–12.

3. Norwitz NG, Soto-Mota A. Case report: Carnivore–ketogenic diet for the treatment of inflammatory bowel disease: a case series of 10 patients. Front Nutr 2024;11:1467475.

4. Chawla S, Silva FT, Medeiros SA, Mekary RA, Radenkovic D. The Effect of Low-Fat and Low-Carbohydrate Diets on Weight Loss and Lipid Levels: A Systematic Review and Meta-Analysis. Nutrients 2020, Vol 12, Page 3774 2020;12:3774.

5. Burén J, Ericsson M, Damasceno N, Sjödin A. A Ketogenic Low-Carbohydrate High-Fat Diet Increases LDL Cholesterol in Healthy, Young, Normal-Weight Women: A Randomized Controlled Feeding Trial. Nutrients 2021;13:814.

6. Norwitz NG, Feldman D, Soto-Mota A, Kalayjian T, Ludwig DS. Elevated LDL Cholesterol with a Carbohydrate-Restricted Diet: Evidence for a “Lean Mass Hyper-Responder” Phenotype. Curr Dev Nutr 2022;6:nzab144.

7. Norwitz NG, Soto-Mota A, Kaplan B, Ludwig DS, Budoff M, Kontush A, Feldman D. The Lipid Energy Model: Reimagining Lipoprotein Function in the Context of Carbohydrate-Restricted Diets. Metabolites 2022, Vol 12, Page 460 2022;12:460.

8. Soto-Mota A, Flores-Jurado Y, Norwitz NG, Feldman D, Pereira MA, Danaei G, Ludwig DS. Increased low-density lipoprotein cholesterol on a low-carbohydrate diet in adults with normal but not high body weight: A meta-analysis. American Journal of Clinical Nutrition 2024;119:740–747.

9. Cooper ID, Sanchez-Pizarro C, Norwitz NG, Feldman D, Kyriakidou Y, Edwards K, Petagine L, Elliot BT, Soto-Mota A. Thyroid markers and body composition predict LDL-cholesterol change in lean healthy women on a ketogenic diet: experimental support for the lipid energy model. Front Endocrinol (Lausanne) 2023;14.

10. Borén J, John Chapman M, Krauss RM, Packard CJ, Bentzon JF, Binder CJ, Daemen MJ, Demer LL, Hegele RA, Nicholls SJ, Nordestgaard BG, Watts GF, Bruckert E, Fazio S, Ference BA, Graham I, Horton JD, Landmesser U, Laufs U, Masana L, Pasterkamp G, Raal FJ, Ray KK, Schunkert H, Taskinen MR, Sluis B van de, Wiklund O, Tokgozoglu L, Catapano AL, Ginsberg HN. Low-density lipoproteins cause atherosclerotic cardiovascular disease: pathophysiological, genetic, and therapeutic insights: a consensus statement from the European Atherosclerosis Society Consensus Panel. Eur Heart J 2020;41:2313–2330.

11. McClelland RL, Jorgensen NW, Budoff M, Blaha MJ, Post WS, Kronmal RA, Bild DE, Shea S, Liu K, Watson KE, Folsom AR, Khera A, Ayers C, Mahabadi AA, Lehmann N, Jöckel KH, Moebus S, Carr JJ, Erbel R, Burke GL. 10-Year Coronary Heart Disease Risk Prediction Using Coronary Artery Calcium and Traditional Risk Factors: Derivation in the MESA (Multi-Ethnic Study of Atherosclerosis) With Validation in the HNR (Heinz Nixdorf Recall) Study and the DHS (Dallas Heart Study). J Am Coll Cardiol 2015;66:1643–1653.

12. Bartlett J, Predazzi IM, Williams SM, Bush WS, Kim Y, Havas S, Toth PP, Fazio S, Miller M. Is isolated low high-density lipoprotein cholesterol a cardiovascular disease risk factor? Circ Cardiovasc Qual Outcomes 2016;9:206–212.

13. Mortensen MB, Caínzos-Achirica M, Steffensen FH, Bøtker HE, Jensen JM, Sand NPR, Maeng M, Bruun JM, Blaha MJ, Sørensen HT, Pareek M, Nasir K, Nørgaard BL. Association of Coronary Plaque With Low-Density Lipoprotein Cholesterol Levels and Rates of Cardiovascular Disease Events Among Symptomatic Adults. JAMA Netw Open. 2022 Feb 1;5(2):e2148139.

14. Fernández-Friera L, Fuster V, López-Melgar B, Oliva B, García-Ruiz JM, Mendiguren J, Bueno H, Pocock S, Ibáñez B, Fernández-Ortiz A, Sanz J. Normal LDL-Cholesterol Levels Are Associated With Subclinical Atherosclerosis in the Absence of Risk Factors. J Am Coll Cardiol 2017;70:2979–2991.

15. Won KB, Park GM, Yang YJ, Ann SH, Kim YG, Yang DH, Kang JW, Lim TH, Kim HK, Choe J, Lee SW, Kim YH, Kim SJ, Lee SG. Independent role of low-density lipoprotein cholesterol in subclinical coronary atherosclerosis in the absence of traditional cardiovascular risk factors. Eur Heart J Cardiovasc Imaging 2019;20:866–872.

16. Javier DAR, Manubolu VS, Norwitz NG, Kinninger A, Aldana-Bitar J, Ghanem A, Ahmad K, Vicuna WD, Hamidi H, Bagheri M, Elsayed T, Villanueva B, Ichikawa K, Flores F, Hamal S, Feldman D, Budoff MJ. The impact of carbohydrate restriction-induced elevations in low-density lipoprotein cholesterol on progression of coronary atherosclerosis: the ketogenic diet trial study design. Coron Artery Dis 2024.

17. Rosendael AR van, Lin FY, Ma X, Hoogen IJ van den, Gianni U, Hussein O Al, Al’Aref SJ, Peña JM, Andreini D, Al-Mallah MH, Budoff MJ, Cademartiri F, Chinnaiyan K, Choi JH, Conte E, Marques H, Araújo Gonçalves P de, Gottlieb I, Hadamitzky M, Leipsic JA, Maffei E, Pontone G, Raff GL, Shin S, Kim YJ, Lee BK, Chun EJ, Sung JM, Lee SE, Berman DS, Virmani R, Samady H, Stone PH, Narula J, Bax JJ, Shaw LJ, Min JK, Chang HJ. Percent atheroma volume: Optimal variable to report whole-heart atherosclerotic plaque burden with coronary CTA, the PARADIGM study. J Cardiovasc Comput Tomogr 2020;14:400–406.

18. Budoff M, Manubolu VS, Kinninger A, Nicholas G. Norwitz P, Feldman D, Thomas R. Wood DBcP, Jonathan Fialkow M, Ricardo Cury M, Theodore Feldman M, Khurram Nasir MM. Carbohydrate Restriction-Induced Elevations in LDL-Cholesterol and Atherosclerosis: The KETO Trial. JACC: Advances 2024;3:101109.

19. van Rosendael AR, Lin FY, van den Hoogen IJ, Ma X, Gianni U, Al Hussein Alawamlh O, Al’Aref SJ, Peña JM, Andreini D, Budoff MJ, Cademartiri F, Chinnaiyan K, Choi JH, Conte E, Marques H, de Araújo Gonçalves P, Gottlieb I, Hadamitzky M, Leipsic J, Maffei E, Pontone G, Raff GL, Shin S, Kim YJ, Lee BK, Chun EJ, Sung JM, Lee SE, Han D, Berman DS, Virmani R, Samady H, Stone P, Narula J, Bax JJ, Shaw LJ, Min JK, Chang HJ. Progression of whole-heart Atherosclerosis by coronary CT and major adverse cardiovascular events. J Cardiovasc Comput Tomogr. 2021;15(4):322–330.

20. Williams MC, Kwiecinski J, Doris M, McElhinney P, D’Souza MS, Cadet S, Adamson PD, Moss AJ, Alam S, Hunter A, Shah ASV, Mills NL, Pawade T, Wang C, Weir McCall J, Bonnici-Mallia M, Murrills C, Roditi G, Beek EJR Van, Shaw LJ, Nicol ED, Berman DS, Slomka PJ, Newby DE, Dweck MR, Dey D. Low-Attenuation Noncalcified Plaque on Coronary Computed Tomography Angiography Predicts Myocardial Infarction: Results from the Multicenter SCOT-HEART Trial (Scottish Computed Tomography of the HEART). Circulation 2020;141:1452–1462.

21. Bär S, Knuuti J, Saraste A, Klén R, Kero T, Nabeta T, Bax JJ, Danad I, Nurmohamed NS, Jukema RA, Knaapen P, Maaniitty T. Derivation and validation of an artificial intelligence-based plaque burden safety cut-off for long-term acute coronary syndrome from coronary computed tomography angiography. Eur Heart J Cardiovasc Imaging 2025;26:1163–1173.

22. Masson W, Siniawski D, Lobo M, Molinero G, Giorgi M, Huerín M. Association between LDL-C, Non HDL-C, and Apolipoprotein B Levels with Coronary Plaque Regression. Arq Bras Cardiol 2015;105:11.

23. Guo Z, Chen G, Ding Y, Wang X, Shan D, Liu Z, Jing J, Chen Y, Yang J. Low-density lipoprotein cholesterol reduction is associated with computed tomography angiography signs of regression and stabilization of coronary plaque: From the TARGET trial. Cardiol Plus 2023;8:269–278.

24. Kambalapalli S, Bhandari M, Punnanithinont N, Iskander B, Khan MA, Budoff M. Bridging Prevention and Imaging: The Influence of Statins on CAC and CCTA Findings. Curr Atheroscler Rep. 2025 Apr 8;27(1):50.

25. Budoff MJ, Kinninger A, Gransar H, Achenbach S, Al-Mallah M, Bax JJ, Berman DS, Cademartiri F, Callister TQ, Chang HJ, Chow BJW, Cury RC, Feuchtner G, Hadamitzky M, Hausleiter J, Kaufmann PA, Leipsic J, Lin FY, Kim YJ, Marques H, Pontone G, Rubinshtein R, Shaw LJ, Villines TC, Min JK. When Does a Calcium Score Equate to Secondary Prevention?: Insights From the Multinational CONFIRM Registry. Cardiovascular Imaging 2023;16:1181–1189.

26. Ahmadi, A, Argulian, E, Leipsic, J. Newby DE, Narula J. From Subclinical Atherosclerosis to Plaque Progression and Acute Coronary Events: *JACC* State-of-the-Art Review. JACC. 2019 Sep, 74 (12) 1608–1617.

27. Budoff MJ, Ellenberg SS, Lewis CE, Mohler ER 3rd, Wenger NK, Bhasin S, Barrett-Connor E, Swerdloff RS, Stephens-Shields A, Cauley JA, Crandall JP, Cunningham GR, Ensrud KE, Gill TM, Matsumoto AM, Molitch ME, Nakanishi R, Nezarat N, Matsumoto S, Hou X, Basaria S, Diem SJ, Wang C, Cifelli D, Snyder PJ. Testosterone Treatment and Coronary Artery Plaque Volume in Older Men With Low Testosterone. JAMA. 2017;317(7):708–716.

28. Narula J, Stuckey TD, Nakazawa G, Ahmadi A, Matsumura M, Petersen K, Mirza S, Ng N, Mullen S, Schaap M, Leipsic J, Rogers C, Taylor CA, Yacoub H, Gupta H, Matsuo H, Rinehart S, Maehara A. Prospective deep learning-based quantitative assessment of coronary plaque by computed tomography angiography compared with intravascular ultrasound: the REVEALPLAQUE study. Eur Heart J Cardiovasc Imaging. 2024 Aug 26;25(9):1287–1295.

29. Rinehart S, Blankstein R, Januzzi JL, McCarthy C, Scherer M, O’Neal WT, Mullen S, Rogers C, Shaw LJ; DECIDE Investigators. Guiding Automated Implementation Strategies for Patients With Atherosclerotic Plaque on Coronary Computed Tomographic Angiography: Rationale and Design of the Artificial Intelligence-DECIDE Study. JACC Cardiovasc Imaging. 2025 Oct;18(10):1107–1115.

30. Ihdayhid AR, Tzimas G, Peterson K, Ng N, Mirza S, Maehara A, Safian RD. Diagnostic Performance of AI-enabled Plaque Quantification from Coronary CT Angiography Compared with Intravascular Ultrasound. Radiol Cardiothorac Imaging. 2024 Dec;6(6):e230312.

