## Supplementary material for "The Impact of Sustained LDL-C Elevation on Plaque Changes: Primary Coronary plaque progression results from the Keto CTA Study": Main Tables

| **Table 1. Baseline Characteristics (N=100)** | | |
| --- | --- | --- |
| Sex (Male) | 59 % | |
| **Ethnicity** | | |
| Asian | 10 % | |
| Hispanic | 7 % | |
| White | 83 % | |
|  | **Mean ± SD** | **Median (Q1-Q3)** |
| Age (years) | 55.3 ± 10.6 | 57.0 (50.8 - 63.0) |
| Ketogenic Diet Duration (days) | 1642.7 ± 913.5 | 1427 (1002 – 1938) |
| Body Mass Index (kg/m^2^) | 22.5 ± 2.7 | 22.3 (20.6 - 24.4) |
| Systolic Blood Pressure (mmHg) | 121.8 ± 21.1 | 120 (112 – 135) |
| Total Cholesterol (mg/dL) | 355.1 ± 89.9 | 338 (301 – 337) |
| LDL-C (mg/dL) | 253.7 ± 84.7 | 237 (202 – 308) |
| HDL-C (mg/dL) | 88.8 ± 20.0 | 88 (74 – 102) |
| Triglycerides (mg/dL) | 66.7 ± 30.4 | 61 (52 – 77) |
| Apolipoprotein B (mg/dL) | 185.0 ± 50.8 | 178 (149 – 214) |
| Lp(a) | 43.4 ± 46.5 | 25.1 (53.7) |
| Noncalcified Plaque Volume (mm^3^) | 32.9 ± 38.5 | 20.4 (6.6 – 48.1) |
| Coronary Artery Calcium Score | 50.3 ± 100.9 | 0 (0 – 54) |
| Percentage Atheroma Volume (%) | 0.6 ± 0.9 | 0.7 (0.2 – 2.0) |
| Low Attenuation Plaque (%) | 0.3 ± 0.8 | 0.06 (0 – 0.3) |
| Total Plaque Volume (mm^3^) | 51.3 ± 71.5 | 24.6 (6.8 – 60.6) |

| **Table 2. Plaque Changes After Follow-Up with QAngio^®^ (pre-registered method) (N=100)** | | | |
| --- | --- | --- | --- |
|  | **Mean (+/- SD)** | | **Median [IQR]** |
| NCPV (mm^3^) | 14.4 ± 26.3 | | 5.6 [15.6] |
| TPV (mm^3^) | 18.7 ± 29.7 | | 6.6 [25.4] |
| PAV (%) | 0.6 ± 0.9 | | 0.2 [0.8] |
| LAP (%) | 0.3 ± 1.4 | | 0 [0.3] |
| **Percentage of participants meeting high-risk plaque cutoffs after follow-up** | | | |
| LAP > 4 % | | 4% | |
| PAV annual change > 1 % | | 21% | |
| **Percentage of participants meeting low-risk plaque cutoffs after follow-up** | | | |
| TPV < 254 mm^3^ | | 94% | |
| PAV < 2.6 % | | 73% | |
| CAC = 0 | | 54% | |
| CAC < 100 | | 80% | |

| **Table 3. Plaque Changes After Follow-Up with Heartflow^®^ (sensitivity analysis) (N=100)** | | | |
| --- | --- | --- | --- |
|  | **Mean (+/- SD)** | | **Median [IQR]** |
| NCPV (mm^3^) | 12.6 ± 46.5 | | 6 [33] |
| TPV (mm^3^) | 14.2 ± 49.8 | | 6 [31.5] |
| PAV (%) | 0.7 ± 2.4 | | 0.3 [1.9] |
| LAP (%) | 0 ± 0.5 | | 0 [0] |
| **Percentage of participants meeting high-risk plaque cutoffs after follow-up** | | | |
| LAP > 4 % | | 0% | |
| PAV annual change > 1 % | | 0% | |
| **Percentage of participants meeting low-risk plaque cutoffs after follow-up.** | | | |
| TPV < 254 mm^3^ | | 88% | |
| PAV < 2.6 % | | 59% | |
