## Supplemental Material for "The Impact of Sustained LDL-C Elevation on Plaque Changes: Primary Coronary plaque progression results from the Keto CTA Study"

| **Supplemental Table 1. Plaque Changes Predictors with QAngio^®^ (N=100)** | | | | | |
| --- | --- | --- | --- | --- | --- |
| **Model** | **β** | **R^2^** | **p-value** | **BF** | **Null Equivalence TOST** |
| ΔNCPV ~ NCVP_Baseline_ | 0.38 | 30% | < 0.001 | < 0.01 | Rejected |
| ΔPAV ~ PAV_Baseline_ | 0.23 | 33% | < 0.001 | < 0.01 | Rejected |
| ΔTPV ~ TVP_Baseline_ | 0.30 | 53% | < 0.001 | < 0.01 | Rejected |
| ΔNCPV ~ CAC_Baseline_ | 0.15 | 32% | < 0.001 | < 0.01 | Rejected |
| ΔPAV ~ CAC_Baseline_ | 0.04 | 24% | < 0.001 | < 0.01 | Accepted |
| ΔTPV ~ CAC_Baseline_ | 0.20 | 45% | < 0.001 | < 0.01 | Rejected |
| ΔNCPV ~ ΔLDL | -0.06 | 2% | 0.07 | 2.0 | Rejected |
| ΔPAV ~ ΔLDL | -0.002 | 4% | 0.02 | 0.75 | Accepted |
| ΔTPV ~ ΔLDL | -0.08 | 3% | 0.05 | 1.5 | Rejected |
| ΔNCPV ~ Days on a Keto Diet | 0.00 | 0% | 0.89 | 9.9 | Accepted |
| ΔPAV ~ Days on a Keto Diet | 0.00 | 0% | 0.98 | 10 | Accepted |
| ΔTPV ~ Days on a Keto Diet | 0.00 | 0% | 0.93 | 9.9 | Accepted |
| ΔNCPV ~ Total LDL Exposure | 0.00 | 0% | 0.87 | 9.9 | Accepted |
| ΔPAV ~ Total LDL Exposure | 0.00 | 0% | 0.72 | 9.4 | Accepted |
| ΔTPV ~ Total LDL Exposure | 0.00 | 0% | 0.93 | 9.9 | Accepted |

BF (Bayes Factors) Values above 1 indicate support for the null-hypothesis, and values below 1 indicate support for the alternative hypothesis. TOST (Two one-sided tests, hypothesis equivalence testing). All equivalence testing used strict bounds for declaring equivalence (CI of 90% for α = 0.01)

| **Supplemental Table 2. Robust Linear Regressions (N=100)** | | |
| --- | --- | --- |
| **Model** | **β** | **p-value** |
| ΔNCPV ~ NCVP_Baseline_ | 0.34 | < 0.001 |
| ΔPAV ~ PAV_Baseline_ | 0.18 | < 0.001 |
| ΔTPV ~ TVP_Baseline_ | 0.27 | < 0.001 |
| ΔNCPV ~ CAC_Baseline_ | 0.15 | < 0.001 |
| ΔPAV ~ CAC_Baseline_ | 0.004 | < 0.001 |
| ΔTPV ~ CAC_Baseline_ | 0.18 | < 0.001 |
| ΔNCPV ~ ΔLDL | -0.02 | 0.12 |
| ΔPAV ~ ΔLDL | -0.00 | 0.20 |
| ΔTPV ~ ΔLDL | -0.03 | 0.16 |
| ΔNCPV ~ Days on a Keto Diet | 0.00 | 0.49 |
| ΔPAV ~ Days on a Keto Diet | 0.00 | 0.43 |
| ΔTPV ~ Days on a Keto Diet | 0.00 | 0.71 |
| ΔNCPV ~ Total LDL Exposure | 0.00 | 0.35 |
| ΔPAV ~ Total LDL Exposure | 0.00 | 0.18 |
| ΔTPV ~ Total LDL Exposure | 0.00 | 0.67 |

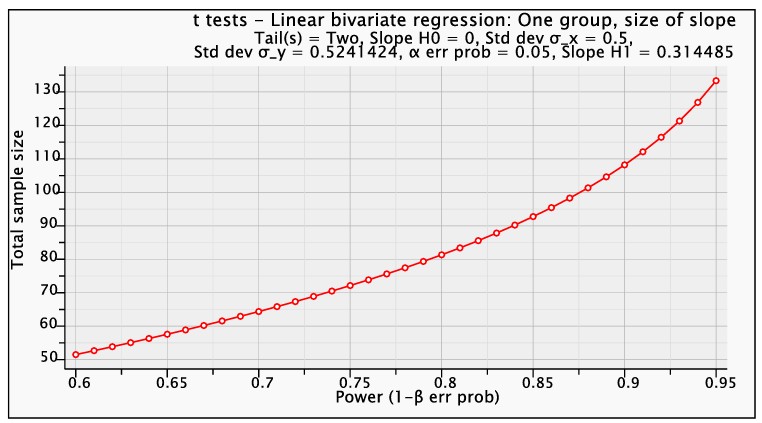
